# Demographic and Clinical Characteristics of the Severe Covid-19 Infections: First Report from Mashhad University of Medical Sciences, Iran

**DOI:** 10.1101/2020.05.20.20108068

**Authors:** Ladan Goshayeshi, Mina Akbari Rad, Robert Bergquist, Abolghasem Allahyari, members of the MUMS Covid-19 Research Team, Benyamin Hoseini

## Abstract

**Background:** Coronavirus Disease 2019 (Covid-19) is expanding worldwide. The characteristics of this infection in patients varies from country to country. To move forward, clinical data on infected patients are needed. Here, we report a comparison between fatalities and recovery of patients with severe Covid-19, based on demographic and clinical characteristics.

**Methods:** Between 5 March and 12 May 2020 in Mashhad, Iran, 1,278 of 4,000 suspected Covid-19 patients were confirmed positive by real-time reverse-transcriptase–polymerase-chain-reaction assay of upper respiratory specimens. We compared the demographic, exposure history and clinical symptoms of 925 survivors and 353 fatal cases with confirmed disease.

**Results:** Mean (SD) age for all confirmed patients was 56.9 (18.7) years, 67.1 (15.9) years in fatal cases and 53.0 (18.3) years in survivors. Multivariable logistic regression analysis showed that the outcome of patients was associated with age (OR = 1.049, P = 0.0001, 95% CI = 1.040-1.057). Despite a high burden of Covid-19 infections in the 30-39 and 40-49 year age groups, most of these (89.6% and 87.2%, respectively) recovered. The median (IQR) duration of hospitalization was 9.0 (6.0-14.0) days. The most prevalent co-morbidities were cardiovascular disorders (21%) and diabetes (16.3%). Dyspnoea (72.7%), cough (68.1%) and fever (63.8%) were the most frequent clinical symptoms. Healthcare workers, of whom two (3%) died, comprised 5.2% of infected-cases. Combination antiviral and antibiotic therapy was used in 43.0% of cases.

**Conclusions:** The characteristics of severe Covid-19 varied substantially between fatal cases and survivors, with diabetes and cardiovascular disorders the most prevalent co-morbidities. In contrast to other studies, there were a higher number of fatalities in younger patients in our settings.

## Background

In late 2019, the city of Wuhan, Hubei Province, China became the centre of an unusual pneumonia-like disease of an unknown cause [1-4]. In December 2019, Chinese scientists realized that this condition was caused by a novel Coronavirus, first called 2019-nCOV [5, 6]. On January 7, 2020, scientists isolated this novel type of Coronavirus from the sputum of these patients, which was on February 11, 2020 classified as Severe Acute Respiratory Syndrome Coronavirus type two (SARS-CoV-2) by the International Committee on Taxonomy of Viruses [6, 7]. On the same date, the World Health Organization (WHO) named the disease caused by this virus coronavirus disease 2019 (Covid-19) in the International Classification of Diseases [6]. As of March 11, 2020 when the Covid-19 outbreak had spread worldwide and reached 118,319 confirmed cases and 4,292 confirmed deaths, WHO declared it a global pandemic [8-10]. The initial source of Covid-19 is still unknown, but it is now confirmed that the first known case of this disease was linked to the Huanan seafood market of Wuhan City, which was closed on January 1, 2020 to control the situation [4, 11].

SARS-CoV-2 is a member of the Coronaviridae family that consists of a single-stranded positive-sense RNA [4]. So far, two human infections by viruses related to Coronaviridae family are known: the Severe Acute Respiratory Syndrome (SARS), caused by SARS-CoV, which emerged in China in 2002 and spread across 37 countries, and the Middle-East Respiratory Syndrome (MERS), caused by MERS-CoV, which was first seen in Saudi Arabia in 2012 [4]. There were two separate outbreaks of SARS in the period 2002-2004, while cases of MERS still occur though only rarely [12]. SARS-COV-2 is genetically related to SARS-CoV with both of them being beta-coronaviruses [13, 14].

The clinical manifestation of Covid-19 varies from asymptomatic infection to severe viral pneumonia with multisystem failure leading to death [3, 15, 16]. The most common clinical manifestations include, but are not limited to, fever, non-productive cough, myalgia, fatigue and dyspnoea. Chest radiography shows bilateral lung infiltrations indicative of pneumonia and laboratory indices include increased erythrocyte sedimentation rate and C-Reactive protein and, lymphopenia [15, 17]. In severe cases, acute respiratory distress syndrome, acute cardiac and kidney injury and shock can occur, which may lead to irreversible organ failure and death [17].

By May 16, 2020, there were 4,602,900 confirmed cases with 307,135 confirmed deaths globally, translating into a case fatality rate (CFR) of 6.67%. At this date, there were 116,635 confirmed cases with 6,902 deaths in Iran resulting in the same CFR (WHO daily report). We aimed to describe the characteristics of patients with Covid-19 in Razavi Khorasan, a province in Iran which has a population of 6.5 million. Herein, we report demographic and clinical characteristics of patients with severe Covid-19 in Mashhad, the second largest city in Iran and capital of Razavi Khorasan.

## Materials and methods

We undertook a retrospective cross-sectional study of consecutive Covid-19 patients diagnosed between 5 March and 12 May 2020 in Mashhad, Iran. Data were collated by the centre for Diseases Prevention and Control of Mashhad University of Medical Sciences. Severe illness usually begins approximately 1 week after the onset of symptoms. Dyspnea is the most common symptom of severe disease and is often accompanied by hypoxemia [6, 17]. Patients with severe Covid-19 should be hospitalized for careful monitoring. In our study all cases were symptomatic and 94% of the cases were inpatients from hospitals affiliated to Mashhad University of Medical Sciences. Thus, we called these cases in this study as sever Covid-19. Among 4,000 mostly severe cases suspected to be infected that had undergone further investigation, 1,278 (32.0%) were confirmed as true Covid-19 cases by real-time reverse-transcriptase-polymerase-chain-reaction (RT-PCR) assay of nasal and\or pharyngeal swabs. Clinical and demographic results of laboratory-confirmed cases were included in a review of exposure history, health outcomes and symptoms. Admission to an intensive care unit (ICU) or the use of mechanical ventilation within this cohort with severe symptoms were the primary end-points of the study. Secondary end-point included duration of hospitalization, history of infection (defined as the time from onset of symptom(s) until discharge from the hospital or death), and outcome (dead or recovered). Fever was defined as axillary temperature >37.5°C. Prescribed medications included lopinavir/ritonavir, antibiotics, and oseltamivir.

## Statistics Analysis

Categorical variables were summarized as frequencies and percentages. The Shapiro–Wilk test was used to evaluate normality of data. Normally distributed data were expressed as mean ± standard deviation (SD), and skewed data as median and interquartile range (IQR). Two-tailed Student’s t and Mann–Whitney U tests were used to compare grouped continuous variable data where appropriate. Chi-square and Fisher’s exact tests were used to perform intergroup and categorical comparisons as appropriate. Logistic regression was performed to multivariable analysis, comparing deceased and recovered patients. Reported p-values of <0.05 were considered statistically significant. SPSS software, version 16 (SPSS Inc., Chicago, IL, USA) was used to analyse the data.

## Results

### Demographic characteristics

Of the 1,278 confirmed Covid-19 patients, 353 (27.6%) died and 925 recovered. Mean (SD) age for confirmed patients was 56.9 (18.7) years, for deceased cases 67.1 (15.9) years and for recovered cases 53.0 (18.3) years. There was a statistically significant association between age and mortality with the mortality rate rising with age (Table 1). Multivariable logistic regression analysis also showed that the outcome of patients was associated with age (OR = 1.049, P = 0.0001, 95% CI = 1.040-1.057), when adjusted for gender, therapeutic approach, body temperature, and duration of hospitalization. Considering age as categorical variable (≤50 vs. >50 years), multivariable logistic regression revealed that odds of death among patients >50 years was more than that among those ≤50 years (OR = 4.7, P = 0.0001, 95% CI = 3.4-6.5). Despite the high frequency of Covid-19 infections accrued in the 30-39 and 40-49 age groups, most of them (130/145 = 89.6 and 170/195 = 87.2%, respectively) recovered (Table 1; Fig. 1). Twenty-six (2.0 %) of all 1.278 confirmed cases were <20 years old, and 28 (2.2%) were >89 years (Fig. 1). Most (793/1,278 = 62%) of the confirmed patients were males, however there was no statistically significant difference in outcomes with reference to gender (Table 1).

**Table 1.** Association between Covid-19 status and outcomes.

| Variable investigated |  | Outcome<br>Number (%) |  | P-value |
| --- | --- | --- | --- | --- |
|  |  | Deceased<br>(n = 353) | Recovered<br>(n = 925) |  |
| Continuous age - mean (SD) |  | 67.1 (15.9) | 53.0 (18.3) | 0.0001 |
| Age (categorical) | 0-9 | 1 (0.3) | 12 (1.3) | 0.0001 |
|  | 10-19 | 1 (0.3) | 12 (1.3) |  |
|  | 20-29 | 4 (1.1) | 66 (7.1) |  |
|  | 30-39 | 15 (4.2) | 130 (14.1) |  |
|  | 40-49 | 25 (7.1) | 170 (18.4) |  |
|  | 50-59 | 47 (13.3) | 193 (20.9) |  |
|  | 60-69 | 104 (29.5) | 165 (17.8) |  |
|  | 70-79 | 74 (21.0) | 102 (11.0) |  |
|  | 80-89 | 62 (17.6) | 67 (7.2) |  |
|  | 90-99 | 19 (5.4) | 8 (0.9) |  |
|  | 100-109 | 1 (0.3) | 0 (0.0) |  |
| Gender | Female | 129 (36.5) | 356 (38.5) | 0.52 |
|  | Male | 224 (63.5) | 569 (61.5) |  |
| Therapeutic approach | Antiviral & antibiotic | 149 (42.2) | 401 (43.4) | 0.08 |
|  | Conservative | 71 (20.1) | 139 (15.0) |  |
|  | Antiviral | 133 (37.7) | 385 (41.6) |  |
| Specific medication used | Lopinavir/ritonavir and antibiotic<sup>a</sup> | 131 (37.1) | 120 (13.0) | NA |
|  | Oseltamivir<sup>b</sup> | 222 (62.9) | 805 (87.0) |  |
| Body temperature (C°) - mean (SD) |  | 37.6 (0.76) | 37.7 (0.73) | 0.09 |
| Hospitalization (days) - median (IQR) |  | 9.0 (6.0-14.0) | 9.0 (6.0-14.0) | 0.18 |

<sup>a</sup>prescribed for severe cases; <sup>b</sup>prescribed for mild cases; NA= Not applicable.

**Fig 1.**
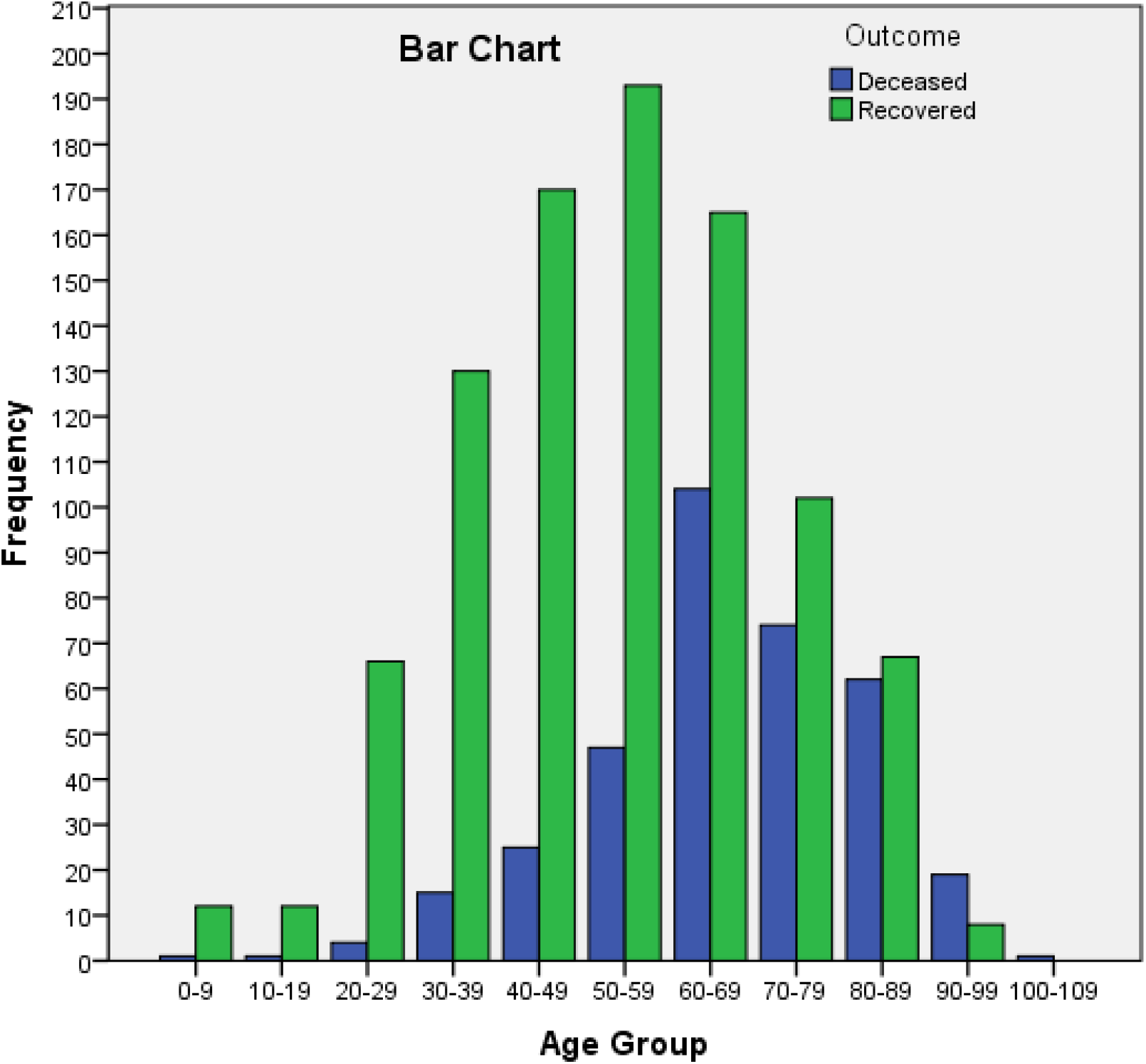
The frequency of the severe Covid-19 infections in different age groups.

### Patient end-points

Of 1,278 patients, 151 (11.8 %) were admitted to ICU of which 70 (46.3%) demised. Furthermore, of 1,278 cases, 499 (39.0%) underwent mechanical ventilation of which 176 (35.3%) demised (Table 2). The median (IQR) duration of hospitalization was 9.0 (6.0-14.0) days, with no significant difference in duration of hospitalization between those that recovered and those who died (Table 1).

**Table 2.** Frequency of co-morbidities and symptoms due to COVID-19 infections.

| Frequency<br>Co-morbidity | Frequency (%) |  |  |
| --- | --- | --- | --- |
|  | Among all infected cases (n= 1,278) | Among deceased cases (n= 353) | Among recovered cases (n= 925) |
| Cardiovascular disorder | 268 (21.0) | 100 (28.3) | 168 (18.2) |
| Diabetes | 208 (16.3) | 81 (22.9) | 127 (13.7) |
| Liver disorders | 45 (3.5) | 18 (5.1) | 27 (2.9) |
| Kidney disorders | 40 (3.1) | 19 (5.4) | 21 (2.3) |
| Chronic resp. disorder | 57 (4.5) | 29 (8.2) | 28 (3.0) |
| Acute resp. distress syndrome (ARDS) | 13 (1.02) | 4 (1.1) | 9 (0.1) |
| Mild pneumonia | 136 (10.64) | 24 (6.8) | 112 (12.1) |
| **Symptom** |  |  |  |
| Fever | 815 (63.8) | 244 (69.1) | 571 (61.7) |
| Cough | 870 (68.1) | 229 (64.9) | 641 (69.3) |
| Dyspnoea | 929 (72.7) | 287 (81.3) | 642 (69.4) |
| General malaise | 247 (19.3) | 61 (17.3) | 186 (20.1) |
| Myalgia | 199 (15.6) | 49 (13.9) | 150 (16.2) |
| Confusion | 87 (6.8) | 41 (11.6) | 46 (5.0) |
| Sore throat | 152 (11.9) | 31 (8.8) | 121 (13.0) |
| Diarrhoea | 47 (3.7) | 11 (3.1) | 36 (3.9) |
| Nausea & vomiting | 113 (8.8) | 27 (7.6) | 86 (9.3) |
| Headache | 114 (8.1) | 24 (6.8) | 90 (9.7) |
| **End-points** |  |  |  |
| ICU admission | 151 (11.8) | 70 (19.8) | 81 (8.7) |
| Using mechanical ventilation | 499 (39.0) | 176 (49.8) | 323 (34.9) |
| **Other** |  |  |  |
| Exposure or visit to hospital within the last 14 days | 131 (10.2) | 38 (10.7) | 93 (10.0) |
| Contact with a respiratory patients in the last 14 days | 616 (48.2) | 126 (35.7) | 490 (53.0) |

### Co-morbidities and symptoms

The most frequent co-morbidities were cardiovascular disorders (268/1,278; 21.0%) and diabetes (208/1,278; 16.3%). Dyspnoea (929, 72.7%), cough (870, 68.1%), and fever (815, 63.8%) were the most frequent clinical symptoms (Table 2). Overall, 1,251/1,278 (97.9%) of the cases had at least one clinical symptom. Compared with survivors, deceased cases showed a higher rate of co-morbidities including cardiovascular disorders (28.3% vs. 18.2%), diabetes (22.9% vs. 13.7%) and chronic respiratory disorders (8.2% vs. 3.0%) (Table 2).

### Therapeutic approach

Most patients (550/1,278; 43%) received antiviral and antibiotic combination therapy, 518/1,278 (40.5%) received antiviral therapies only, and 210/1,278 (16.5%) did not receive antiviral or antibiotic therapy. No significant differences in the treatment approach between patients that died and those that recovered were observed (Table 1). Mortality rates associated with lopinavir/ritonavir and antibiotic combination therapy and oseltamivir therapy were 37.1% and 62.9%, respectively (Table 1).

Of the 1,278 confirmed cases, 67 (5.2%) were healthcare workers, of whom 2 (3.0%) died. Thirty-one (46.3%) of these healthcare workers were presumed to have been infected through occupational exposure to infected patients. Cough (77.6%), fever (68.7) and dyspnoea (56.7%) were the commonest symptoms in healthcare workers with Covid-19. Six (9.0%) had comorbidities (cardiovascular disorders, n=3; diabetes, n=3). Seven (10.4%) of the healthcare workers required mechanical ventilation and 6 were admitted to ICU.

## Discussion

This is the first study from Mashhad, Iran, to report the characteristics of patients with severe confirmed Covid-19. Increasing age was associated with an increased risk of mortality. The most frequent co-morbidities were cardiovascular disorders and diabetes, and the most frequent symptoms were dyspnoea, cough, and fever. The highest burden of disease clustered in the 50-59 and 60-69 year age groups, while the highest CFR was patients aged 80-89 years. These findings are in line with previous reports [6, 18].

Mean (SD) age of Iranian Covid-19 patients in our study was 56.9 (18.7) years, which is similar to other studies [17, 19, 20], but older than that reported by Guan et al. [2] in china who reported a median age (IQR) of 47.0 (35.0 - 58.0) years, with 55.1% of their cases were between 15-49 years, and younger than the median of 64 years reported in other studies [4, 21]. Although, the most frequent Covid-19 infection occurred in the 50-59 and 60-69 age groups in our study, the frequency of infection was also considerable in the 30-39 and 40-49 year age groups. The high frequency of Covid-19 infection in these age groups may be due to the low median age (30 years) of the Iranian population and/or due to inclusion of mainly working-age population in this age group [22]. It seems that the frequency of Covid-19 infections rises considerably from age group of 50-59 years [23, 24]. In contrast to findings in China and Italy [6, 25], we found a considerably higher proportion of fatalities in the age groups panning the range from 20-50 years. This high proportion of fatalities in young people may be due to differences in life expectancy of different countries [26]. For lower- and middle-income countries, the proportion of deaths among younger age groups appears substantially larger. Another reason for this high proportion of fatalities in young people may be due to unknown co-morbidities of these people. Elderly people with known co-morbidities may take more care of themselves than young people with unknown co-morbidities. Also, it may be due to shortage of ICU that caused some patients were mechanically ventilated in ordinary wards, i.e. outside ICU.

In our study, the prevalence of the Covid-19 infection was higher among men (62%) compared with the reports of China (49.3% - 54.3%) [17, 21]. However, our finding of male predominance in severe cases was similar to reports from Italy (60%) [27], and the United States (63%) [4].

Huang et al [18] reported a CFR of 15% among hospitalized Covid-19 cases in China. According to the WHO daily report, the crude CFR was 5.6% in China by May 10, 2020. At the beginning of the Covid-19 outbreak in Italy, the crude CFR was 11.8% (12,430/105,792) [28], but this decreased as the number of cases increased. In our study, RT-PCR testing was only used for symptomatic cases, including more severely ill patients at a high risk of mortality. This selection bias, and the fact that our study describes outcomes in confirmed cases during a comparatively short window of time, limits our ability to arrive at accurate estimates of CFR relating Covid-19 in our setting. Long-term screening of all population at risk should facilitate reporting of more reliable CFR estimates in future studies. Estimation of the infection fatality rate (IFR) would also be of interest, but would require reliable antibody detection assay and large population-based samples.

According to WHO interim clinical guidance (Rev. March 20, 2020) [29], fever, caught, fatigue and anorexia are the most common signs of Covid-19 and our findings comply with this. However, in our study dyspnoea (72.7%) was twice as prevalent compared to WHO estimates (31-40%) [29]. Prevalence of dyspnoea in our study, was similar to that from United States, however [4, 30]. The prevalence of dyspnoea in China complies with the WHO guidelines [19, 21, 31], although they may also be slightly higher [32]. The WHO should update the information on symptoms and signs of Covid-19 based on new global data, pointing out that there may be differences with respect to region.

Cardiovascular disorders (21%) and diabetes (16.3%) were the most prevalent co-morbidities among Covid-19 patients in our study. Previous studies have reported diverse prevalence rates for cardiovascular disorders (11-45%) and diabetes (13-35%) [4, 19, 30, 33]. Meta-analyses report pooled prevalence of 8.4% and 12% for cardiovascular disorders, and 8.0% to 9.7% for diabetes [34, 35]. Assessing the association between patients’ awareness of Covid-19 and risk factors for severe disease and adherence to prevention protocols should be evaluated in future studies.

The fatality rate among healthcare workers was 3% in our study, considerably higher than that reported in China (0.33%) [36]. This may be due to a shortage of personal protective equipment (PPE) at the early stages of the Covid-19 pandemic in Iran. Future studies should focus on the impact that availability of PPE had on CFR among healthcare workers in Iran, and other countries.

Huang et al [37] reported that 32% of patients were admitted to ICU, whereas in our study only 11.8% received ICU care. The fatality rate among patients treated in ICU was 46.3% in our study, lower than that (78%) reported by Zhou et al. [6]. All patients admitted to ICU received broad-spectrum antibiotics and antiviral medications, a combination that may have contributed to the lower mortality rate in our setting. Importantly, the use if the RT-PCR testing of all ICU-admitted patients strengthened the reliability of CFR estimates of this category of patients.

To our knowledge, this is the first study to address the characteristics of Covid-19 patients confirmed by RT-PCR in Mashhad, Iran. However, it has limitations, chiefly in that the RT-PCR testing approach focused on cases with severe symptoms, most of which were hospitalized. We analysed data from patients treated in facilities served by the Mashhad University of Medical Sciences, and so did not cover all areas of Razavi Khorasan, although most patients requiring hospitalization are routinely referred to this city. Despite the above-mentioned limitations, we feel that our findings are instructive and will inform future research, both in Iran and internationally.

In summary, the frequency of Covid-19 fatalities rose considerably from 30-39 years, with a higher number of fatalities in younger patients than in international studies. Fever, cough, and dyspnoea were common symptoms among hospitalized patients with Covid-19. Diabetes and cardiovascular disorders were the most prevalent co-morbidities, as observed in other studies.

## Data Availability

All data will be available upon request by permission of the corresponding author and Research Ethics Committee of Mashhad University of Medical Sciences.

## List of abbreviations

Covid-19: Coronavirus Disease 2019
SARS-CoV-2: severe acute respiratory syndrome coronavirus 2
SARS: severe acute respiratory syndrome
ICU: intensive care unit
RT-PCR: real-time reverse-transcriptase polymerase chain reaction

## Declarations

*Ethics approval*. This study was approved by the Ethics Committee of the Mashhad University of Medical Science (Ethics code: IR.MUMS.REC.1398.308).

*Consent for publication*. Not applicable.

*Availability of data and materials*. The datasets used and/or analysed during the current study are available from the corresponding author on reasonable request.

*Competing interests*. The authors declare that they have no competing interests.

*Funding*. This work was supported by Mashhad University of Medical Sciences (grant number of 981800).

*Authors’ contributions*. BH, RB designed the study design. All authors contributed in data gathering and interpretation of the results. LG, MAR, BH wrote the first draft of the manuscript. RB edited final version of the manuscript. All authors read, commented and approved the final manuscript.

## Acknowledgment

We would like to thank all personnel of Mashhad University of Medical Sciences for their supports.

